# Phenome-wide association of acne susceptibility loci highlights shared and unique biology

**DOI:** 10.1101/2023.06.19.23291559

**Authors:** Minsoo Kim, Carol Cheng

## Abstract

The latest genome-wide association studies (GWAS) comprising 20,165 acne cases and 595,231 controls have successfully identified 42 genomic regions associated with increased risk of acne. Yet, it remains unclear the extent to which these individual acne susceptibility loci contribute to other disorders. To begin to identify shared and unique biology underlying acne pathogenesis, we conducted phenome-wide association (PheWAS) of the 42 acne loci across 110 phenotypes with large-scale, well-powered GWAS results that broadly span autoimmune, endocrine, psychiatric, cardiovascular, dermatologic conditions, and cancer, as well as serum and urine biomarkers. Much of the acne loci did not share GWAS signals with any of 110 phenotypes, suggesting that there may be acne-specific biology in these genomic regions. Conversely, we also identified shared genetic effects at several loci with testosterone, sex hormone binding globulin (SHBG), non-albumin protein levels, liver enzymes (AST, AST/ALT ratio), and blood cell phenotypes (MCV, MCH, eosinophil count), some of which recapitulate known acne pathophysiology. Overall, this work highlights shared and unique genetic effects of individual acne GWAS loci via phenome-wide investigation. All GWAS summary statistics used herein and code to reproduce bioinformatic data analyses are publicly available.

## RESULTS AND DISCUSSION

Acne vulgaris is a highly heritable and debilitating, common inflammatory skin disorder that is characterized by comedones, papules, pustules, nodules, and cystic lesions. Despite its tremendous contribution to public health burden worldwide, there have been few advances in the treatment of acne since 1982, due in large part to the lack of novel, robust therapeutic targets. Fortunately, the latest genome-wide association studies (GWAS) (Mitchell et al. 2022) comprising 20,165 acne cases and 595,231 controls have successfully identified 42 genomic loci associated with increased risk of acne (Figure 1), which brings hope that genetics can provide novel insights into underlying acne mechanisms and identify new biological pathways for intervention (Common et al. 2019; van Steensel 2019).

**Figure 1:**
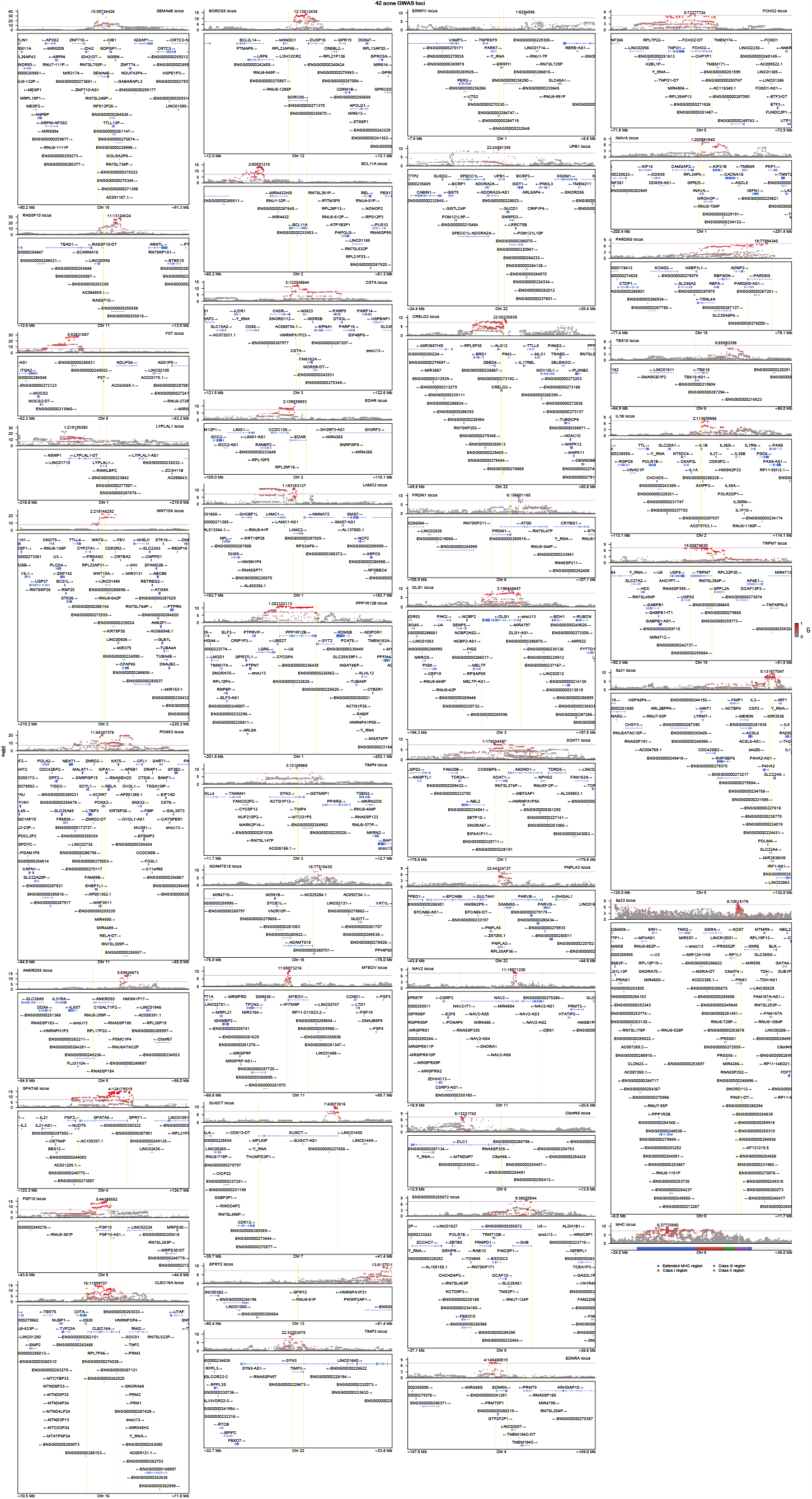
LocuZoom plots of 42 acne GWAS loci. Each locus is named after a gene that is the closest to the most significant SNP (i.e. index SNP) from either 5’ or 3’ end. Shown are ±0.5 Mb windows of corresponding gene start and end sites. Index SNPs are labeled as purple diamond and LD between other SNPs are displayed with the intensity of red color. LD is calculated with individuals of European ancestry in the 1000 Genomes Project reference panel. Gene annotation is based on GENCODE v39 and genomic coordinates are based on the GRCh37 human genome build. Purple line denotes genome-wide significance threshold (*P* = 5e-8), and yellow lines denote gene start and end sites. The 39 non-complicated GWAS loci are visualized in the order of the strength of their association from top to bottom and left to right. For example, the most significant GWAS signal for acne resides within the 15q26 region with multiple independent signals within the *SEMA4B* gene. The three loci that reside in complex regions of the human genome with extended LD (i.e. 5q31, 8p23, and MHC) are also shown in the bottom right corner.

Previous work (Mitchell et al. 2022) has revealed substantial genetic overlap between acne and several disorders, such as Crohn’s disease, breast cancer, and schizophrenia. However, little is known about the extent to which these individual acne loci contributes to other disorders. Hence, to begin to identify shared and unique biology underlying acne pathogenesis, we conducted phenome-wide association (PheWAS) of the 42 acne loci across 110 phenotypes with large-scale, well-powered GWAS results that broadly span autoimmune, endocrine, psychiatric, cardiovascular, dermatologic conditions, and cancer, as well as serum and urine biomarkers (Supplementary Figures S1-42).

Of the 42 acne loci, three resided within complex regions of the human genome, namely 5q31, 6p21, and 8p23 (Figure 1), which are characterized by long-range linkage disequilibrium (LD) and known to harbor complex structural variation and/or copy number variation (Anderson et al. 2010). These regions were in general highly pleiotropic—for example, the major histocompatibility complex (MHC) region on 6p21 essentially harbored a significant association for every phenotype (Supplementary Figure S42). Similarly, 8p23 harbored a significant association for nearly half of phenotypes (53 of 110) (Supplementary Figure S41). Since complex LD patterns can complicate comparison of GWAS signals, we ignore these regions hereafter and focus on the remaining 39 loci.

On visual inspection (Figure 1), it was clear that at least 9 (*SEMA4B, RASSF10, LYPLAL1, PCNX3, CLEC16A, SUGCT, UPB1, PRDM1, PNPLA3*) of the 39 loci harbored multiple independent association signals, suggesting that there may be more GWAS signals than previously recognized (Mitchell et al. 2022). We note that conditional analysis using summary statistics (Yang et al. 2012) was not possible in this study due to the lack of relevant information (e.g. effect size estimates and their standard errors). Given that testing for colocalization of GWAS signals using existing approaches is difficult when multiple independent signals are present (Wallace 2021), we instead utilized visual inspection and manually compared GWAS signals across the 39 acne loci. While most loci harbored distinct GWAS signals, 9 loci showed strong evidence of colocalization (Figure 2), which we discuss below.

**Figure 2:**
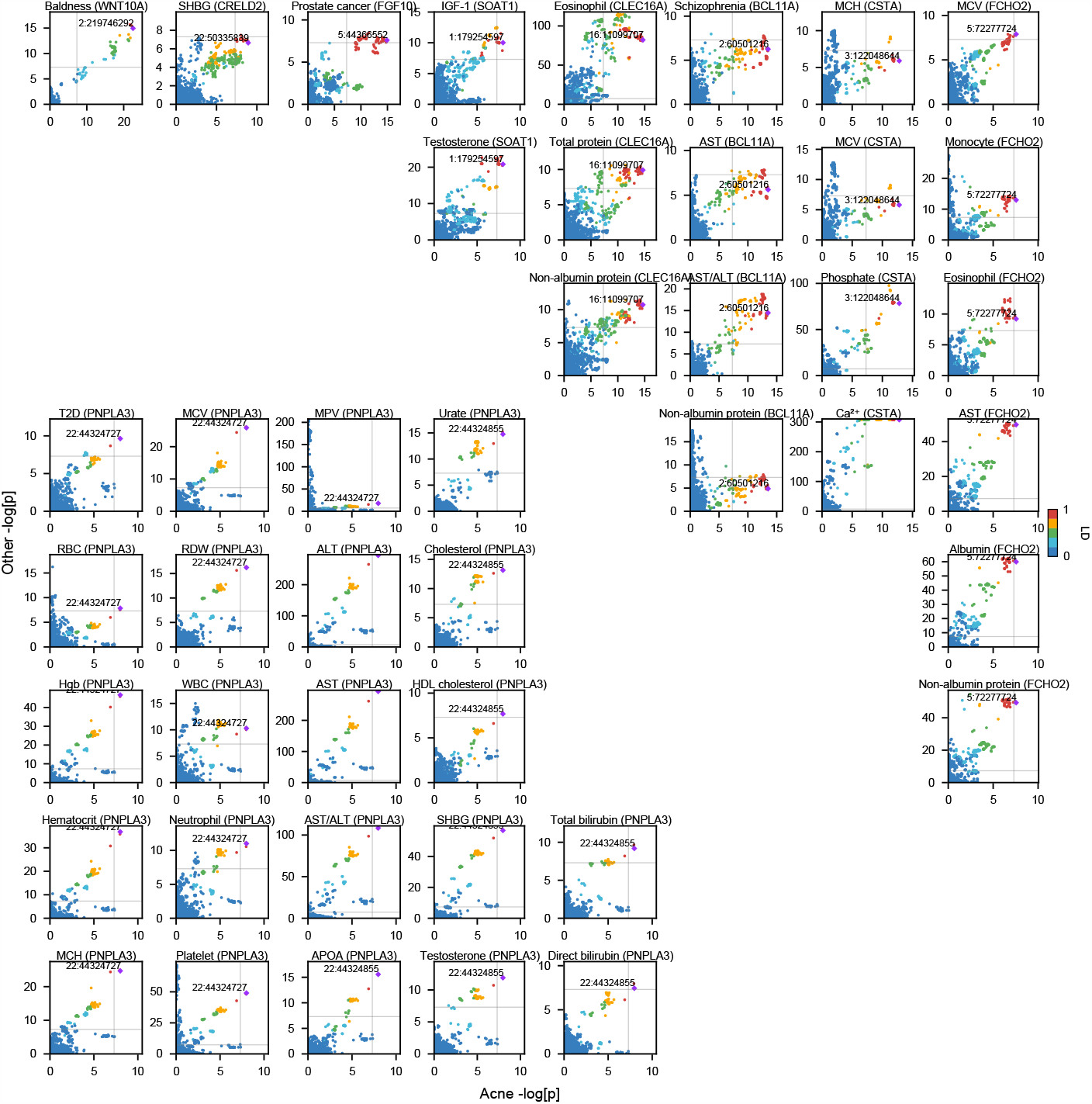
Colocalization of acne GWAS signals in 9 loci. Shown is the strength of association as -log_10_*P* for acne and other relevant GWAS results. Index SNPs for acne GWAS are labeled as purple diamond and LD between other SNPs are displayed. LD is calculated with individuals of European ancestry in the 1000 Genomes Project reference panel. Genomic coordinates are based on the GRCh37 human genome build. Gray line denotes genome-wide significance threshold (*P* = 5e-8). Baldness (male-pattern baldness), Eosinophil (eosinophil count), MCH (mean corpuscular hemoglobin), MCV (mean corpuscular volume), Monocyte (monocyte count), MPV (mean platelet volume), Neutrophil (neutrophil count), RBC (RBC count), RDW (red cell distribution width), SHBG (sex hormone binding globulin), T2D (type II diabetes), WBC (WBC count).

Genetic variation within the *PNPLA3* gene colocalized with the largest number of phenotypes (22 of 110) (Figure 2, Supplementary Figure S29). The LD block here tightly hugged the 5’ ends of *PNPLA3* and *PARVB* genes (Figure 1), and there seemed to be at least two independent association signals within this locus, one of which colocalized with 22 phenotypes, including diabetes, liver enzymes, cholesterol, bilirubin levels, testosterone, sex hormone binding globulin (SHBG), and several blood cell phenotypes. Meanwhile, *FCHO2* locus harbored the second most number of colocalization with phenotypes like albumin and non-albumin protein levels, eosinophil and monocyte counts (Figure 2, Supplementary Figure S34). Its index SNP resided within the gene close to the 5’ end, and the LD block hugged the 5’ ends of *TNPO1* and *TMEM171* genes. Of note, *FCHO2* is found to be constrained with pLI = 1.

*CSTA* and *BCL11A* had the third most number of colocalization, each with 4 phenotypes. *CSTA* locus harbored a fairly large LD block, and its index SNP was within the gene body close to the 5’ end. It shared a GWAS signal with calcium, phosphate levels, mean corpuscular volume (MCV), and mean corpuscular hemoglobin (MHC) (Figure 2, Supplementary Figure S13). Of note, *CTSA* is associated with peeling skin syndromes. On the other hand, *BCL11A* shared a GWAS signal with schizophrenia, non-albumin protein, and liver enzymes levels (Figure 2, Supplementary Figure S12). It may be worthwhile to note that the GWAS signal for this locus resided within an intergenic region, unlike in previous examples.

*CLEC16A* is another acne-associated locus that seemingly harbored multiple independent GWAS signals with the first signal in the *CLEC16A* and the second in the *RMI2* gene bodies. The first signal colocalized with that of eosinophil count and serum protein levels (Figure 2, Supplementary Figure S10). Next, *SOAT1* locus harbored a fairly large LD block with its index SNP close to the 5’ end of *SOAT1* gene. This GWAS signal colocalized with that of IGF-1 and testosterone levels (Figure 2, Supplementary Figure S28). Finally, *FGF10, CRELD2*, and *WNT10A* each shared a GWAS signal with a single phenotype. For *FGF10*, the index SNP resided within the gene, and its association pattern matched that of prostate cancer (Figure 2, Supplementary Figure S9). *FGF10* is also found to be constrained with pLI = 0.94. Meanwhile, for *CRELD2*, the LD block hugged the gene boundaries of *BRD1* and *CRELD2* with the index SNP being in close proximity to the 3’ end of *CRELD2* gene. Acne and SHBG shared a GWAS signal in this locus (Figure 2, Supplementary Figure S25). For *WNT10A*, its index SNP resided near the 5’ end of *WNT10A* and its association pattern matched that of male-pattern baldness (Figure 2, Supplementary Figure S5). Of note, *WNT10A* is associated with tooth morphology and ectodermal dysplasia (Xu et al. 2017).

In summary, we systematically examined acne GWAS loci phenome-wide to assess the degree of shared genetic effects and shared biology. Acne shared the largest number of GWAS signals with AST, MCV, and non-albumin protein levels at three, while it shared two GWAS signals with testosterone, SHBG levels, AST/ALT ratio, MCH, and eosinophil counts. Shared genetic effects with testosterone and SHBG makes intuitive sense given that acne is associated with androgen excess. On the contrary, relations between acne and liver enzymes, blood cell phenotypes, protein levels remain unclear, although we do note that liver function is routinely monitored for patients on isotretinoin. Whether these phenotypes with shared genetics have a causal effect on the development of acne remains to be investigated in the future. We further note that there were several loci where it was difficult to determine sharing of GWAS signals. In these loci, there were genetic variants that reached genome-wide significance in both acne and another GWAS, but without straightforward pattern of colocalization. This may be due to LD mismatch between different studies or presence of multiple causal variants and warrants further follow-up. The remaining loci did not share GWAS signals with any of 110 phenotypes, suggesting that there may be acne-specific biology in these genomic regions. However, we caution the readers in interpreting this finding, as we have only looked at 110 phenotypes and the number of phenotypes with shared biology may increase in the future when looking at genetic association results of more acne-relevant phenotypes.

## Data Availability

No new data have been generated as part of this study. All GWAS summary statistics used for data analyses and data visualization herein are publicly available.

https://github.com/mmkim1210/acne-genetics

## DATA AVAILABILITY

No new data have been generated as part of this study. All GWAS summary statistics used for data analyses and data visualization herein are publicly available. For URLs to download them, please see https://github.com/mmkim1210/GeneticsMakie.jl/blob/master/src/gwas.jl. The code used to perform bioinformatic analyses are available at https://github.com/mmkim1210/acne-genetics. All plots were generated using Makie.jl (Danisch and Krumbiegel 2021) and GeneticsMakie.jl (Kim et al. 2022).

## ACKNOWLEDGEMENTS

This work was supported by the UCLA-Caltech Medical Scientist Training Program (T32GM008042).

## AUTHOR CONTRIBUTIONS

Conceptualization: MK; Formal Analysis: MK; Interpretation: MK, CC; Writing: MK.

## FIGURES

**Supplementary Figures S1-42: Phenome-scale LocusZoom plots for 42 acne GWAS loci**. GWAS results for 110 complex phenotypes are shown, which span autoimmune, endocrine, psychiatric, cardiovascular disorders, and cancer. Index SNPs for phenotypes harboring GWAS hits are labeled and corresponding linkage disequilibrium (LD) between other SNPs are displayed with the intensity of red color. Purple line denotes genome-wide significance (*P* = 5e-8), and yellow lines denote gene start and end sites for the cognate gene, which is defined as the closest gene to the index SNP either from 5’ or 3’ end. ±1 Mb window around the cognate gene is plotted, except for those in complex regions of the human genome (i.e. 5q31, 8p23, and MHC), for which the entire region is visualized. Note that some -log_10_*P* values are clamped to 308, since their *P* values cannot be represented by the smallest floating-point number. ADHD (attention-deficit/hyperactivity disorder), ALS (amyotrophic lateral sclerosis), AMD (age-related macular degeneration), BD (bipolar disorder), CAD (coronary artery disease), CKD (chronic kidney disease), DBP (diastolic blood pressure), IBD (inflammatory bowel disease), PCOS (polycystic ovarian syndrome), RBC (red blood cell), SBP (systolic blood pressure), SHBG (sex hormone binding globulin), SCZ (schizophrenia).

